# The association between age and high-intensity treatment in traumatic brain injury patients: a CENTER-TBI study

**DOI:** 10.1101/2024.06.25.24309449

**Authors:** Ernest van Veen, Erwin J. O. Kompanje, Mathieu van der Jagt, Ana Mikolić, Giuseppe Citerio, Nino Stocchetti, Diederik Gommers, David K. Menon, Andrew I. R. Maas, Hester F. Lingsma, the CENTER-TBI investigators and participants

**Affiliations:** Department of Public Health, Erasmus University Medical Center, Rotterdam, the Netherlands; Department of Intensive Care Adults, Erasmus MC - University Medical Center, Rotterdam, the Netherlands; Department of Medical Ethics and Philosophy of Medicine, Erasmus University Medical Center, Rotterdam, the Netherlands; School of Medicine and Surgery, University of Milano-Bicocca, Milano, Italy; Fondazione IRCCS San Gerardo dei Tintori, Monza, Italy; Department of Physiopathology and Transplantation, Milan University, Milan, Italy; Neuro ICU Fondazione IRCCS Cà Granda Ospedale Maggiore Policlinico Milano, Milan, Italy; Department of Anaesthesia, University of Cambridge, Cambridge, United Kingdom; Department of Neurosurgery, Antwerp University Hospital and University of Antwerp, Edegem, Belgium

**Keywords:** Traumatic brain injury, critical care, intensive care units, elderly

## Abstract

**Introduction:** Older age is associated with worse outcome after traumatic brain injury (TBI). Whether this association is entirely driven by frailty, or clinicians’ reluctance to give specific treatments to older patients is unclear. Therefore, we aimed to confirm the association between age and worse outcome, and to assess the association between age and received high-intensity treatment (HIT).

**Methods:** We included TBI patients aged 16 and older from the CENTER-TBI study. The association between age and HIT, and between age and outcome (the Glasgow Outcome Scale Extended (GOSE) at 6 months), was analyzed using multivariable ordinal and logistic regression respectively. In the overall cohort, HIT was defined as receiving emergency intracranial surgery, or ICU admission. In the subset of patients admitted to the ICU, HIT was defined as receiving metabolic suppression, intensive hypocapnia, hypothermia below 35 °C, decompressive craniectomy, or intracranial surgery not scheduled on admission. We adjusted for pre-injury health, injury severity (Glasgow Coma Scale (GCS) motor score and pupillary reactivity at baseline; major extracranial injury (MEI); Marshall CT classification), and in the subset of ICU patients for the median ICP before receiving any HIT.

**Results:** In total, 4349 patients were included. Of these, 1999 patients (46%) were admitted to the ICU. The median age was 51 years. Every ten-year increase in age for patients over 65, was associated with worse outcome (OR = 0.6, CI = 0.5 – 0.7, OR in the subset of ICU patients = 0.5, CI = 0.3 – 0.5). Furthermore, every ten-year increase in age for patients over 65 was associated with a lower likelihood of receiving emergency intracranial surgery (OR = 0.4, CI = 0.3 – 0.6), and ICU admission (OR = 0.6, CI = 0.5 – 0.8). Similarly, in the subset of ICU patients, every ten-year increase in age for patients over 65, was associated with a lower likelihood of receiving neuromuscular blockade (OR = 0.6, CI = 0.4 – 0.9), intensive hypocapnia (OR = 0.2, CI = 0.1 – 0.9), decompressive craniectomy (OR = 0.4, CI = 0.2 – 0.8), and intracranial surgery (OR = 0.5, CI = 0.3 – 0.8).

**Conclusion:** Older patients have poorer outcome, and were less likely to receive high-intensity treatments, independent of patient and injury characteristics. Clinicians should not withhold high-intensity treatments solely based on older age. Educating clinicians about this delicate topic, and performing further comparative effectiveness research focusing on older patients may improve diagnosis, treatments, and understanding of TBI outcomes in this group.

## INTRODUCTION

Traumatic brain injury (TBI) is a serious societal problem and a tremendous burden for patients and their families (1). In recent decades, TBI occurrence shifted towards the older age groups (≥65 years) (2), presenting with a low-energy fall (2). This shift in TBI epidemiology is relevant since older age is associated with worse outcomes after TBI (3–9).

Despite this shift, older adults with TBI are often excluded from research (10), even though scientific evidence is important for the creation of guidelines. Without these guidelines, clinicians have to balance risks and benefits of treatments individually. This may in turn lead to differences in outcome (11), as is warned for in, among other disciplines, older adults with cancer (12), and rheumatoid arthritis (13).

Examples of differences in management between older and younger adults with TBI, are the influence of age on the decision to perform neurosurgical procedures (14), or to implement high-intensity treatment (HIT) (15, 16). Limitations of these studies include the sample size, and lack of adjustment for intracranial pressure (ICP). Pessimistic assumptions (based on these limitations) could increase the risk of a self-fulfilling prophecy, negatively impacting (long-term) outcomes for older adults (15).

To add to the scarce literature on older adults with TBI, we aimed to confirm the association between increasing age and worse outcome, and to assess the association between age and received HIT.

## METHODS

### Study population

The Collaborative European NeuroTrauma Effectiveness Research in TBI (CENTER-TBI, registered at clinicaltrials.gov NCT02210221) is a prospective cohort study conducted in 63 centers from 18 countries across Europe and Israel between 2014 and 2017. Patients were included if they arrived at the hospital within 24 hours after injury with a clinical diagnosis of TBI, and if patients had an indication for a head computed tomography (CT) scan. Patients with a severe preexisting neurological disorder that would confound outcome assessment, were excluded. For each center ethics approval was acquired and consent for participation was obtained from all patients or their proxies. The general methods and aims of the CENTER-TBI study were previously described (17, 18).

For this study, we included all patients aged 16 or older. We also specifically studied the subgroup of patients admitted to the ICU. We extracted data on demographics, (pre)injury, admission, imaging, monitoring, treatment, and outcome characteristics.

### Treatments

We selected treatment variables based on clinical considerations. These treatment variables were described in the e-CRF exactly as written below, and were scored (if applicable) once daily. In the entire cohort (patients admitted to the ER, ward, or ICU), high-intensity treatment (HIT) was defined as one or more of the following treatments:

- ICU admission (yes/no).
- Emergency intracranial operation (having received a craniotomy, decompressive craniectomy, depressed skull fracture or other intracranial procedures) (yes/no).

In patients admitted to the ICU, HIT was defined as having received at least one of the following treatments:

- Metabolic suppression for ICP control with high dose barbiturates or propofol during the ICU stay (yes/no).
- Neuromuscular blockade (paralysis) during ICU stay (yes/no).
- Intensive hypocapnia [PaCO2 < 4.0 kPa (30 mmHg)] during ICU stay (yes/no).
- Hypothermic treatment (only treatment, not accidental temperatures) below 35°C during ICU stay (yes/no).
- Decompressive craniectomy (yes/no).
- Intracranial operation for progressive mass lesion, not scheduled on admission (yes/no).

### Outcomes

We used the Glasgow Outcome Scale-Extended (GOSE) as the outcome measure (19). GOSE is an eight-point outcome scale that was measured six months after injury: (1) dead; (2) vegetative state; (3) lower severe disability; (4) upper severe disability; (5) lower moderate disability; (6) upper moderate disability; (7) lower good recovery; and (8) upper good recovery. Categories 2 and 3 were combined, resulting in a seven-point ordinal scale.

### Statistical analyses

#### Descriptive statistics

Baseline characteristics were presented as median values with interquartile ranges (IQRs) for continuous variables and as frequencies and percentages for categorical variables. We compared characteristics between age groups (i.e. age 65 or over vs. age 65 or under) for the entire and ICU cohort. We used the Pearson χ2 test for categorical variables and the independent t-test or Mann– Whitney U-Test for continuous variables to test for differences between these groups.

#### Baseline variables

We collapsed categories 3 and 4 of the American Society of Anesthesiologists (ASA) Physical Status classification because category 4 had <10 patients. Second, we collapsed categories V and VI of the Marshall CT classification as grading V and VI could not be differentiated on central review as the raters were unaware of (intent to) surgery. Last, to compare groups at baseline we calculated the expected probability of mortality and unfavorable outcome using the International Mission for Prognosis and Analysis of Clinical Trials in TBI (IMPACT) core model (8).

#### Regression

We used ordinal and logistic regression, with GOSE and treatments as dependent variables to assess:

1. To confirm if older age is associated with worse outcome (GOSE) (3–9).
2. To assess if older age is associated with a lower probability to receive HIT (15).

All models were adjusted for age; sex; cause of injury (road traffic accident, fall, or other); American Society of Anesthesiologists Physical Status (ASAPS) classification (healthy patient, patients with mild systemic disease, patients with severe systemic disease); pre-injury use of anticoagulants or platelet inhibitors; pre-injury use of beta-blockers; GCS motor score at baseline; pupillary reactivity at baseline; major extracranial injury (MEI); Marshall CT classification. Models for the subset of patients admitted to the ICU, were additionally adjusted for the median ICP scores measured before a patient received any HIT in the ICU.

We analyzed age as a continuous variable and allowed for non-linear effects by estimating different effects for patients over and under 65. As a sensitivity analysis we estimated different effects for patients over and under 50, based on several important longitudinal studies of aging, and a recent systematic review on geriatric patients with TBI (3, 20, 21). Furthermore, as a sensitivity analysis, based on clinical judgment, we estimated different effects for patients over and under 80.

Associations between age and GOSE were expressed as Odds Ratios (ORs) with 95% confidence intervals (CIs). The ordinal logistic regression estimates a common OR for overall health state transitions within the GOSE.

All statistical analyses were performed in R studio (22). Multiple imputation was used to handle missing values, using the mice package in R (23). Data were accessed using ‘Neurobot’ (http://neurobot.incf.org; RRID: SCR_01700), vs 3.0 (data freeze: October 2021).

## RESULTS

### Characteristics study cohort – (*Table 1*-2)

The entire study cohort consisted of 4349 patients. The median age was 51 years old (IQR 32-67), and the main cause of injury was an incidental fall (1955, 46%) (Table 1). The ICU cohort consisted of 1999 patients. The median age was 51 years old (IQR 31-66), and the main cause of injury was a road traffic accident (887, 45%) (Table 2).

**Table 1.** Baseline characteristics entire cohort.

| Table 1. Baseline characteristics entire cohort |  |  |  |  |  |
| --- | --- | --- | --- | --- | --- |
|  | All<br>n = 4349 | ≤65 years old<br>n = 3156 | >65 years old<br>n = 1193 | p-<br>value | Missing |
| **Patient characteristics** |  |  |  |  |  |
| Age (median [IQR]) | 51.00<br>[32.00, 67.00] | 42.00<br>[27.00, 54.00] | 75.00<br>[70.00, 80.00] | <0.001 | 0.0 |
| Sex male (%) | 2926 (67.3) | 2226 (70.5) | 700 (58.7) | <0.001 | 0.0 |
| Preinjury ASAPS classification (%) |  |  |  | <0.001 | 3.1 |
| A normal healthy patient | 2352 (55.8) | 2118 (69.3) | 234 (20.2) |  |  |
| A patient with a mild systemic disease | 1401 (33.3) | 771 (25.2) | 630 (54.5) |  |  |
| A patient with a severe systemic disease (that is a constant threat to life)** | 460 (10.9) | 167 (5.5) | 293 (25.3) |  |  |
| Use of anticoagulants or platelet inhibitors (%) | 738 (17.6) | 182 (6.0) | 556 (49.0) | <0.001 | 3.8 |
| Use of betablockers (%) | 435 (10.5) | 125 (4.1) | 310 (28.1) | <0.001 | 4.8 |
| ICU admission (%) | 2055 (47.3) | 1530 (48.5) | 525 (44.0) | 0.009 | 0.0 |
| **Baseline characteristics** |  |  |  |  |  |
| Injury cause (%) |  |  |  | <0.001 | 2.8 |
| Road traffic incident | 1619 (38.3) | 1355 (44.2) | 264 (22.7) |  |  |
| Incidental fall | 1955 (46.2) | 1148 (37.5) | 807 (69.3) |  |  |
| Other | 654 (15.5) | 560 (18.3) | 94 (8.1) |  |  |
| GCS motor baseline (%) |  |  |  | 0.041 | 2.3 |
| 1 | 647 (15.2) | 488 (15.8) | 159 (13.6) |  |  |
| 2 | 77 (1.8) | 54 (1.8) | 23 (2.0) |  |  |
| 3 | 79 (1.9) | 68 (2.2) | 11 (0.9) |  |  |
| 4 | 151 (3.6) | 109 (3.5) | 42 (3.6) |  |  |
| 5 | 420 (9.9) | 305 (9.9) | 115 (9.9) |  |  |
| 6 | 2874 (67.7) | 2057 (66.8) | 817 (70.0) |  |  |
| Pupils baseline (%) |  |  |  | 0.412 | 5.9 |
| Both reactive | 3654 (89.3) | 2667 (89.6) | 987 (88.4) |  |  |
| One reactive | 162 (4.0) | 117 (3.9) | 45 (4.0) |  |  |
| Both unreactive | 277 (6.8) | 192 (6.5) | 85 (7.6) |  |  |
| Major Extracranial Injury (%) | 1533 (35.2) | 1180 (37.4) | 353 (29.6) | <0.001 | 0.0 |
| **CT characteristics** |  |  |  |  |  |
| Marshall CT classification (%) |  |  |  | <0.001 | 7.7 |
| I | 1545 (38.5) | 1188 (40.9) | 357 (32.3) |  |  |
| II | 1434 (35.7) | 1053 (36.2) | 381 (34.5) |  |  |
| III | 152 (3.8) | 128 (4.4) | 24 (2.2) |  |  |
| IV | 33 (0.8) | 28 (1.0) | 5 (0.5) |  |  |
| V/VI*** | 849 (21.2) | 511 (17.6) | 338 (30.6) |  |  |
| Epidural hematoma present on CT (%) | 445 (10.2) | 385 (12.2) | 60 (5.0) | <0.001 | 0.0 |
| Acute subdural hematoma present on CT (%) | 1173 (27.0) | 751 (23.8) | 422 (35.4) | <0.001 | 0.0 |
| Acute subarachnoid hemorrhage present on CT (%) | 1858 (42.7) | 1276 (40.4) | 582 (48.8) | <0.001 | 0.0 |
| Midline shift present on CT (%) | 476 (20.5) | 297 (9.4) | 179 (15.0) | <0.001 | 0.0 |
| **Surgical interventions** |  |  |  |  |  |
| Emergency intracranial surgery during hospital stay (%) | 506 (11.7) | 380 (12.2) | 126 (10.6) | 0.186 | 0.9 |
| Emergency extracranial surgery during hospital stay (%) | 337 (7.8) | 286 (9.2) | 51 (4.3) | <0.001 | 0.9 |
| Intracranial surgery during hospital stay (%) | 859 (24.3) | 642 (25.4) | 217 (21.6) | 0.020 | 18.7 |
| Extracranial surgery during hospital stay (%) | 714 (20.2) | 587 (23.2) | 127 (12.6) | <0.001 | 18.7 |
| **Outcomes** |  |  |  |  |  |
| Length of hospital stay in days (median [IQR]) | 4.52 [0.99, 14.99] | 4.49 [0.96, 15.74] | 4.66 [1.05, 13.47] | 0.965 | 2.6 |
| IMPACT core probability of mortality (%)**** | 19 [12, 24] | 15 [0.09, 0.19] | 0.24 [0.24, 0.24] | <0.001 | 6.6 |
| IMPACT core probability of unfavorable outcome (%)**** | 40 [26, 47] | 33 [21, 40] | 47 [47, 47] | <0.001 | 6.6 |
| Glasgow Outcome Scale Extended (%) |  |  |  | <0.001 | 15.8 |
| Dead | 470 (12.8) | 200 (7.6) | 270 (26.4) |  |  |
| Vegetative state/lower severe disability | 314 (8.6) | 199 (7.5) | 115 (11.2) |  |  |
| Upper severe disability | 174 (4.8) | 120 (4.5) | 54 (5.3) |  |  |
| Lower moderate disability | 339 (9.3) | 295 (11.2) | 44 (4.3) |  |  |
| Upper moderate disability | 383 (10.5) | 333 (12.6) | 50 (4.9) |  |  |
| Lower good recovery | 658 (18.0) | 491 (18.6) | 167 (16.3) |  |  |
| Upper good recovery | 1325 (36.2) | 1002 (38.0) | 323 (31.6) |  |  |

Abbreviations: ASAPS = American Society of Anesthesiologists Physical Status; CT = computed tomography; GCS = Glasgow coma scale; ICP = intracranial pressure; IMPACT = International Mission for Prognosis and Analysis of Clinical Trials in TBI; IQR = interquartile range; TBI = traumatic brain injury
\* P-values are a comparison between patients below versus over 65 years old.
\*\* Category 3 and 4 of the ASAPS classification were collapsed since category 4 had <50 patients.
\*\*\* Grading V and VI of the Marshall CT classification were collapsed since raters were not aware of (intent for) surgery and could thus not differentiate between grade V and VI
\*\*\*\* To calculate the probability of mortality and unfavorable outcome, we used the International Mission for Prognosis and Analysis of Clinical Trials in TBI (IMPACT) core model (8).

**Table 2.**
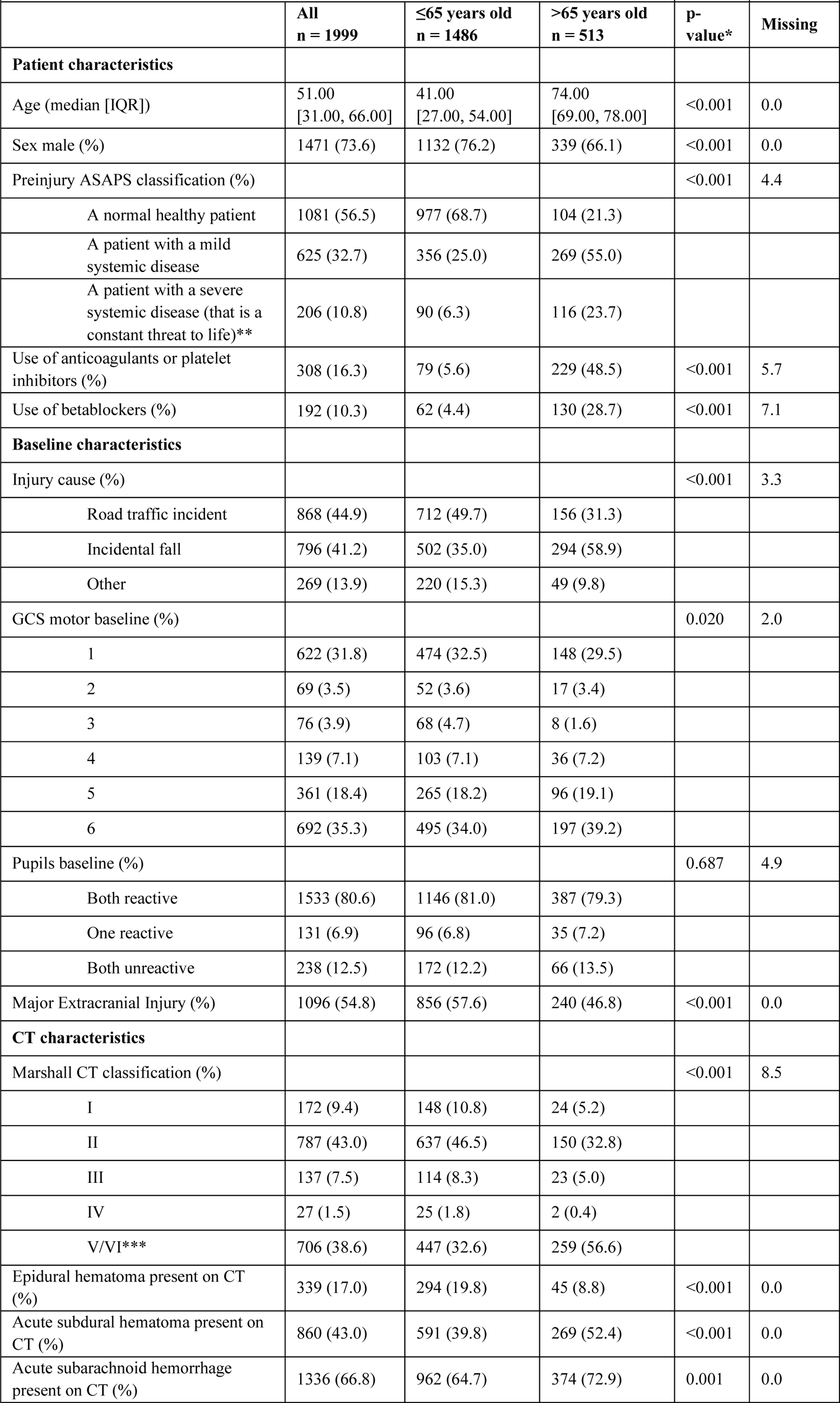
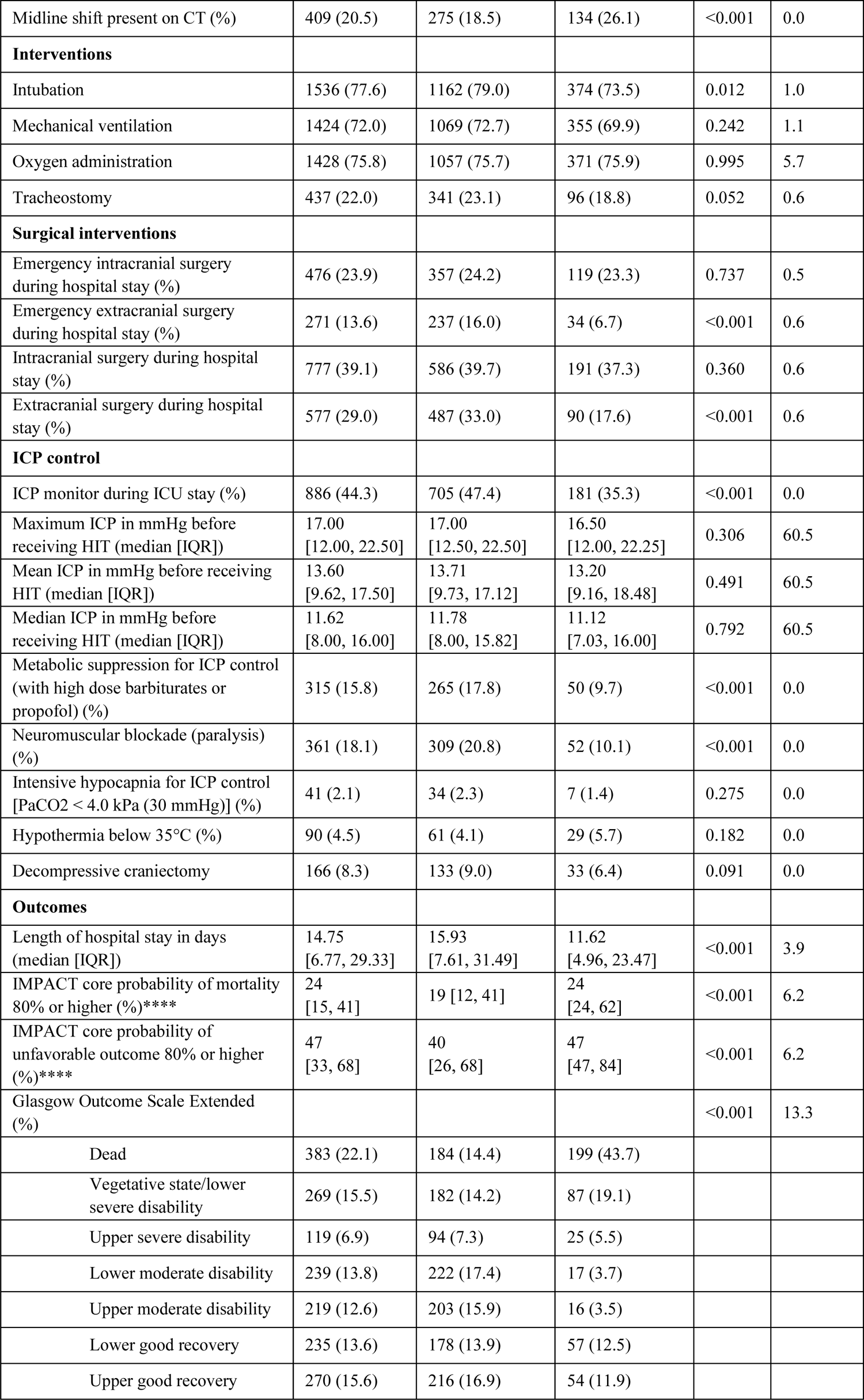
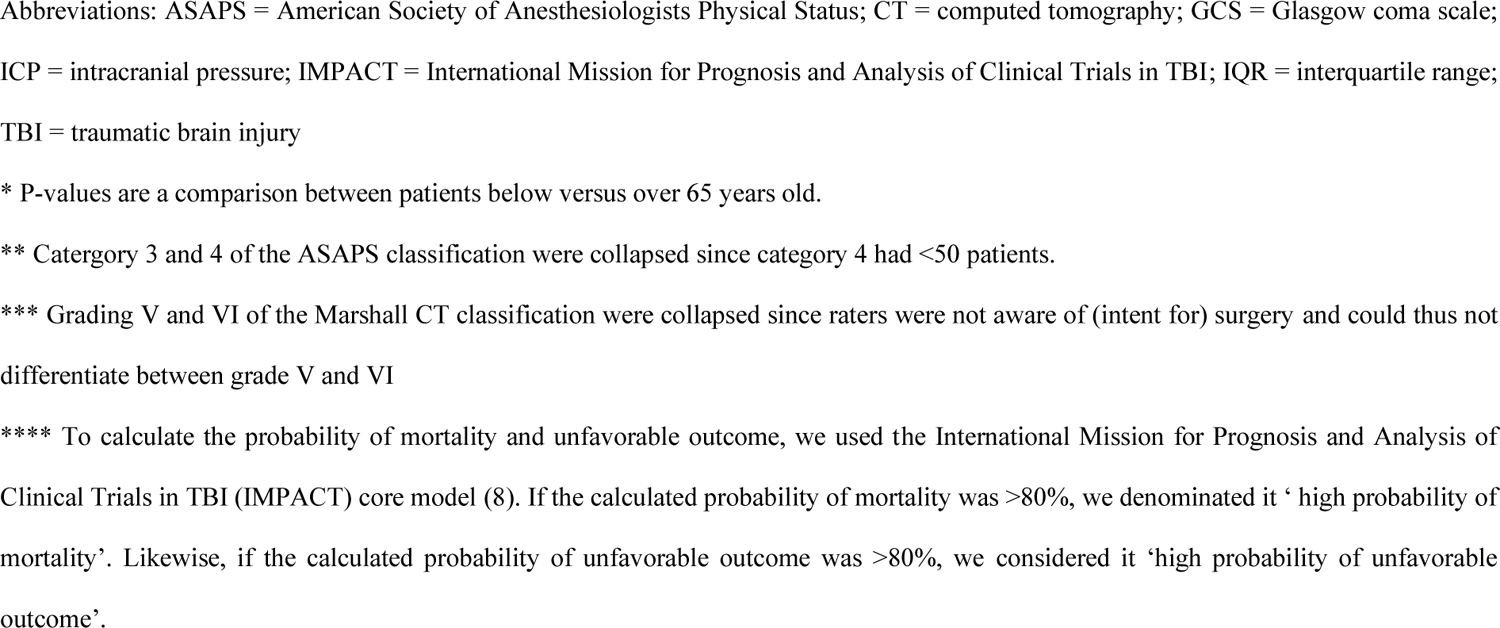
Baseline characteristics ICU cohort.

| Table 2. Baseline characteristics ICU cohort |  |  |  |  |  |
| --- | --- | --- | --- | --- | --- |
|  | All<br>n = 1999 | ≤65 years old<br>n = 1486 | >65 years old<br>n = 513 | p-<br>value* | Missing |
| **Patient characteristics** |  |  |  |  |  |
| Age (median [IQR]) | 51.00<br>[31.00, 66.00] | 41.00<br>[27.00, 54.00] | 74.00<br>[69.00, 78.00] | <0.001 | 0.0 |
| Sex male (%) | 1471 (73.6) | 1132 (76.2) | 339 (66.1) | <0.001 | 0.0 |
| Preinjury ASAPS classification (%) |  |  |  | <0.001 | 4.4 |
| A normal healthy patient | 1081 (56.5) | 977 (68.7) | 104 (21.3) |  |  |
| A patient with a mild systemic disease | 625 (32.7) | 356 (25.0) | 269 (55.0) |  |  |
| A patient with a severe systemic disease (that is a constant threat to life)** | 206 (10.8) | 90 (6.3) | 116 (23.7) |  |  |
| Use of anticoagulants or platelet inhibitors (%) | 308 (16.3) | 79 (5.6) | 229 (48.5) | <0.001 | 5.7 |
| Use of betablockers (%) | 192 (10.3) | 62 (4.4) | 130 (28.7) | <0.001 | 7.1 |
| **Baseline characteristics** |  |  |  |  |  |
| Injury cause (%) |  |  |  | <0.001 | 3.3 |
| Road traffic incident | 868 (44.9) | 712 (49.7) | 156 (31.3) |  |  |
| Incidental fall | 796 (41.2) | 502 (35.0) | 294 (58.9) |  |  |
| Other | 269 (13.9) | 220 (15.3) | 49 (9.8) |  |  |
| GCS motor baseline (%) |  |  |  | 0.020 | 2.0 |
| 1 | 622 (31.8) | 474 (32.5) | 148 (29.5) |  |  |
| 2 | 69 (3.5) | 52 (3.6) | 17 (3.4) |  |  |
| 3 | 76 (3.9) | 68 (4.7) | 8 (1.6) |  |  |
| 4 | 139 (7.1) | 103 (7.1) | 36 (7.2) |  |  |
| 5 | 361 (18.4) | 265 (18.2) | 96 (19.1) |  |  |
| 6 | 692 (35.3) | 495 (34.0) | 197 (39.2) |  |  |
| Pupils baseline (%) |  |  |  | 0.687 | 4.9 |
| Both reactive | 1533 (80.6) | 1146 (81.0) | 387 (79.3) |  |  |
| One reactive | 131 (6.9) | 96 (6.8) | 35 (7.2) |  |  |
| Both unreactive | 238 (12.5) | 172 (12.2) | 66 (13.5) |  |  |
| Major Extracranial Injury (%) | 1096 (54.8) | 856 (57.6) | 240 (46.8) | <0.001 | 0.0 |
| **CT characteristics** |  |  |  |  |  |
| Marshall CT classification (%) |  |  |  | <0.001 | 8.5 |
| I | 172 (9.4) | 148 (10.8) | 24 (5.2) |  |  |
| II | 787 (43.0) | 637 (46.5) | 150 (32.8) |  |  |
| III | 137 (7.5) | 114 (8.3) | 23 (5.0) |  |  |
| IV | 27 (1.5) | 25 (1.8) | 2 (0.4) |  |  |
| V/VI*** | 706 (38.6) | 447 (32.6) | 259 (56.6) |  |  |
| Epidural hematoma present on CT (%) | 339 (17.0) | 294 (19.8) | 45 (8.8) | <0.001 | 0.0 |
| Acute subdural hematoma present on CT (%) | 860 (43.0) | 591 (39.8) | 269 (52.4) | <0.001 | 0.0 |
| Acute subarachnoid hemorrhage present on CT (%) | 1336 (66.8) | 962 (64.7) | 374 (72.9) | 0.001 | 0.0 |
| Midline shift present on CT (%) | 409 (20.5) | 275 (18.5) | 134 (26.1) | <0.001 | 0.0 |
| **Interventions** |  |  |  |  |  |
| Intubation | 1536 (77.6) | 1162 (79.0) | 374 (73.5) | 0.012 | 1.0 |
| Mechanical ventilation | 1424 (72.0) | 1069 (72.7) | 355 (69.9) | 0.242 | 1.1 |
| Oxygen administration | 1428 (75.8) | 1057 (75.7) | 371 (75.9) | 0.995 | 5.7 |
| Tracheostomy | 437 (22.0) | 341 (23.1) | 96 (18.8) | 0.052 | 0.6 |
| **Surgical interventions** |  |  |  |  |  |
| Emergency intracranial surgery during hospital stay (%) | 476 (23.9) | 357 (24.2) | 119 (23.3) | 0.737 | 0.5 |
| Emergency extracranial surgery during hospital stay (%) | 271 (13.6) | 237 (16.0) | 34 (6.7) | <0.001 | 0.6 |
| Intracranial surgery during hospital stay (%) | 777 (39.1) | 586 (39.7) | 191 (37.3) | 0.360 | 0.6 |
| Extracranial surgery during hospital stay (%) | 577 (29.0) | 487 (33.0) | 90 (17.6) | <0.001 | 0.6 |
| **ICP control** |  |  |  |  |  |
| ICP monitor during ICU stay (%) | 886 (44.3) | 705 (47.4) | 181 (35.3) | <0.001 | 0.0 |
| Maximum ICP in mmHg before receiving HIT (median [IQR]) | 17.00<br>[12.00, 22.50] | 17.00<br>[12.50, 22.50] | 16.50<br>[12.00, 22.25] | 0.306 | 60.5 |
| Mean ICP in mmHg before receiving HIT (median [IQR]) | 13.60<br>[9.62, 17.50] | 13.71<br>[9.73, 17.12] | 13.20<br>[9.16, 18.48] | 0.491 | 60.5 |
| Median ICP in mmHg before receiving HIT (median [IQR]) | 11.62<br>[8.00, 16.00] | 11.78<br>[8.00, 15.82] | 11.12<br>[7.03, 16.00] | 0.792 | 60.5 |
| Metabolic suppression for ICP control (with high dose barbiturates or propofol) (%) | 315 (15.8) | 265 (17.8) | 50 (9.7) | <0.001 | 0.0 |
| Neuromuscular blockade (paralysis) (%) | 361 (18.1) | 309 (20.8) | 52 (10.1) | <0.001 | 0.0 |
| Intensive hypocapnia for ICP control [PaCO<sub>2</sub> < 4.0 kPa (30 mmHg)] (%) | 41 (2.1) | 34 (2.3) | 7 (1.4) | 0.275 | 0.0 |
| Hypothermia below 35°C (%) | 90 (4.5) | 61 (4.1) | 29 (5.7) | 0.182 | 0.0 |
| Decompressive craniectomy | 166 (8.3) | 133 (9.0) | 33 (6.4) | 0.091 | 0.0 |
| **Outcomes** |  |  |  |  |  |
| Length of hospital stay in days (median [IQR]) | 14.75<br>[6.77, 29.33] | 15.93<br>[7.61, 31.49] | 11.62<br>[4.96, 23.47] | <0.001 | 3.9 |
| IMPACT core probability of mortality 80% or higher (%)**** | 24<br>[15, 41] | 19 [12, 41] | 24<br>[24, 62] | <0.001 | 6.2 |
| IMPACT core probability of unfavorable outcome 80% or higher (%)**** | 47<br>[33, 68] | 40<br>[26, 68] | 47<br>[47, 84] | <0.001 | 6.2 |
| Glasgow Outcome Scale Extended (%) |  |  |  | <0.001 | 13.3 |
| Dead | 383 (22.1) | 184 (14.4) | 199 (43.7) |  |  |
| Vegetative state/lower severe disability | 269 (15.5) | 182 (14.2) | 87 (19.1) |  |  |
| Upper severe disability | 119 (6.9) | 94 (7.3) | 25 (5.5) |  |  |
| Lower moderate disability | 239 (13.8) | 222 (17.4) | 17 (3.7) |  |  |
| Upper moderate disability | 219 (12.6) | 203 (15.9) | 16 (3.5) |  |  |
| Lower good recovery | 235 (13.6) | 178 (13.9) | 57 (12.5) |  |  |
| Upper good recovery | 270 (15.6) | 216 (16.9) | 54 (11.9) |  |  |

Abbreviations: ASAPS = American Society of Anesthesiologists Physical Status; CT = computed tomography; GCS = Glasgow coma scale; ICP = intracranial pressure; IMPACT = International Mission for Prognosis and Analysis of Clinical Trials in TBI; IQR = interquartile range; TBI = traumatic brain injury
\* P-values are a comparison between patients below versus over 65 years old.
\*\* Category 3 and 4 of the ASAPS classification were collapsed since category 4 had <50 patients.
\*\*\* Grading V and VI of the Marshall CT classification were collapsed since raters were not aware of (intent for) surgery and could thus not differentiate between grade V and VI
\*\*\*\* To calculate the probability of mortality and unfavorable outcome, we used the International Mission for Prognosis and Analysis of Clinical Trials in TBI (IMPACT) core model (8). If the calculated probability of mortality was >80%, we denominated it 'high probability of mortality'. Likewise, if the calculated probability of unfavorable outcome was >80%, we considered it 'high probability of unfavorable outcome'.

### Baseline characteristics ≤65 years versus >65 years – entire cohort (*Table 1*)

Of the 4349 patients, 1139 (26.2%) were >65 years old. Patients >65 were less often male (58.7% versus 70.5%), and the most prevalent cause of injury in patients >65 was a fall (69.3%), whereas it was a road traffic incident in patients ≤65 (44.2%). Patients >65 more often had a mild or severe systemic disease pre-injury (79.8% versus 30.7%), and more often used medication such as anticoagulants or platelet inhibitors (49% versus 6%), and beta-blockers (28.1% versus 4.1%). Patients >65 more often had a GCS motor score of 6 (70% versus 66.8%), more often had a Marshall CT classification of V or VI (30.6% versus 17.6%), but less often had MEI (29.6% versus 37.4%). Patients >65 and ≤65 had a similar length of hospital stay of generally 5 days. Six months after their hospital stay, patients >65 were more often deceased than younger patients (26.4% versus 7.6% respectively) (Table 1).

### Baseline characteristics ≤65 years versus >65 years – ICU cohort (*Table 2*)

Of the 1999 patients, 513 (25.7%) were >65 years old. Patients >65 were less often male (66.1% versus 76.2%), and the most prevalent cause of injury in patients >65 was a fall (58.9%), whereas it was a road traffic incident in patients ≤65 (49.7%). Patients >65 more often had a mild or severe systemic disease pre-injury (78.7% versus 31.3%), and more often used medication such as anticoagulants or platelet inhibitors (48.5 % versus 5.6%), and beta-blockers (28.7% versus 4.4%). Patients >65 less often had a GCS motor score of 1 (29.5% versus 49.7%), but more often had a Marshall CT classification of V or VI (56.6% versus 32.6%). MEI was less prevalent in patients >65 (46.8% versus 57.6%). Patients >65 were less often intubated (73.5% versus 79.0%), and less often had an ICP monitor during their ICU stay (35.3% versus 47.4%), with a median ICP before receiving any ICP lowering therapies of 11.12 mmHg compared to 11.78 mmHg in patients ≤65. Patients >65 generally had a shorter length of hospital stay of 12 days compared to 16 days for their younger counterparts. Six months after their hospital stay, patients >65 were more often deceased than patients ≤65 (44.1% versus 14.3% respectively) (Table 2).

#### Association of age with GOSE (Table 3-5, Figure 1-3)

Increasing age was independently associated with worse GOSE for patients in the entire cohort (OR per 10 years if over 65 = 0.62, CI = 0.54 – 0.70) and in the ICU cohort (OR per 10 years if over 65 = 0.48, CI = 0.33 – 0.52) (Table 3, Figure 1-3). This was also the case for patients under 65 in the entire cohort (OR per 10 years = 0.90, CI = 0.84 – 0.91), and in the ICU cohort (OR per 10 years = 0.84, CI = 0.80 – 0.91) (Table 3). Comparable associations were found in the sensitivity analyses.

**Figure 1.**
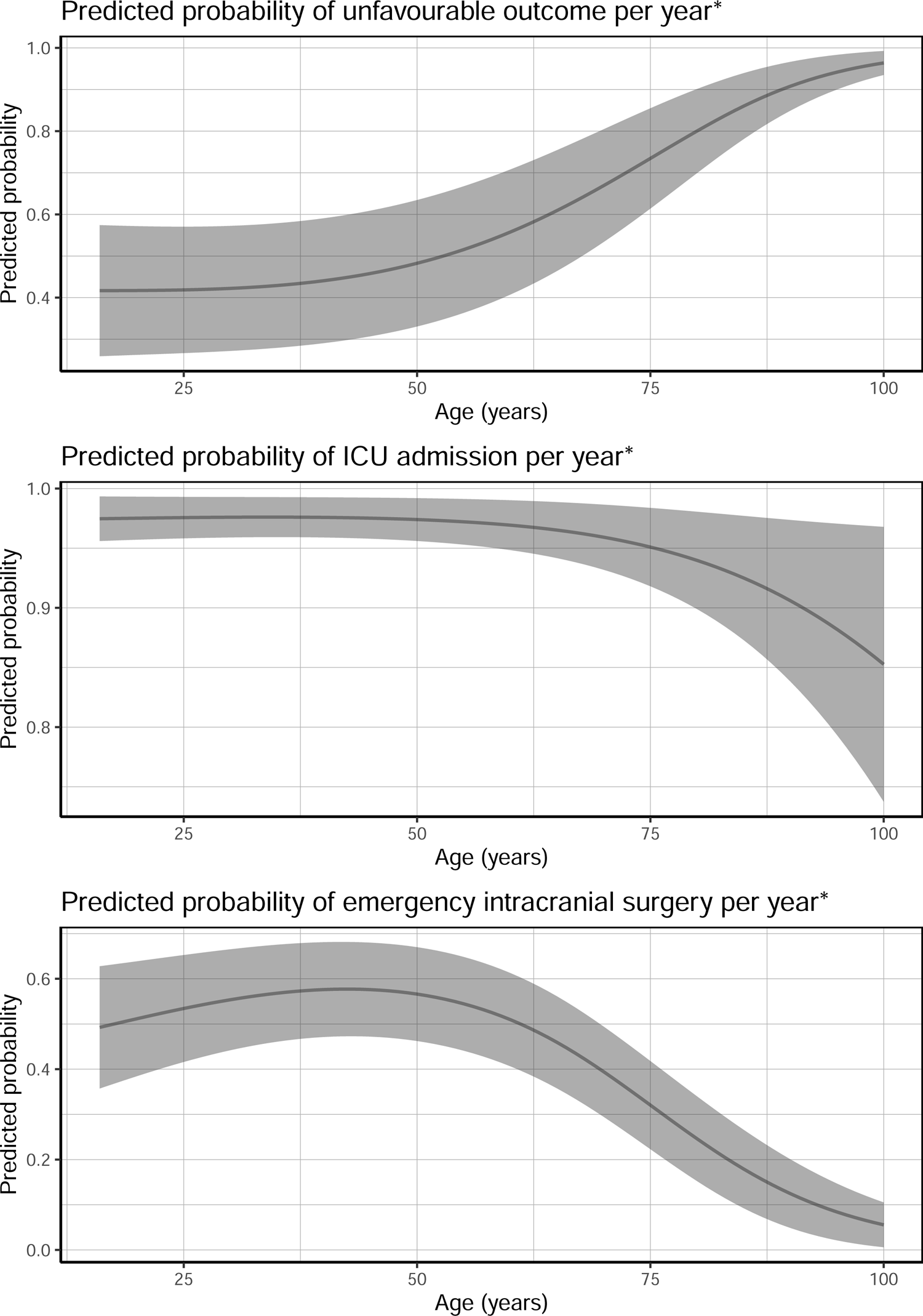
Predicted probabilities of unfavorable outcome, ICU admission and emergency intracranial surgery

**Figure 2.**
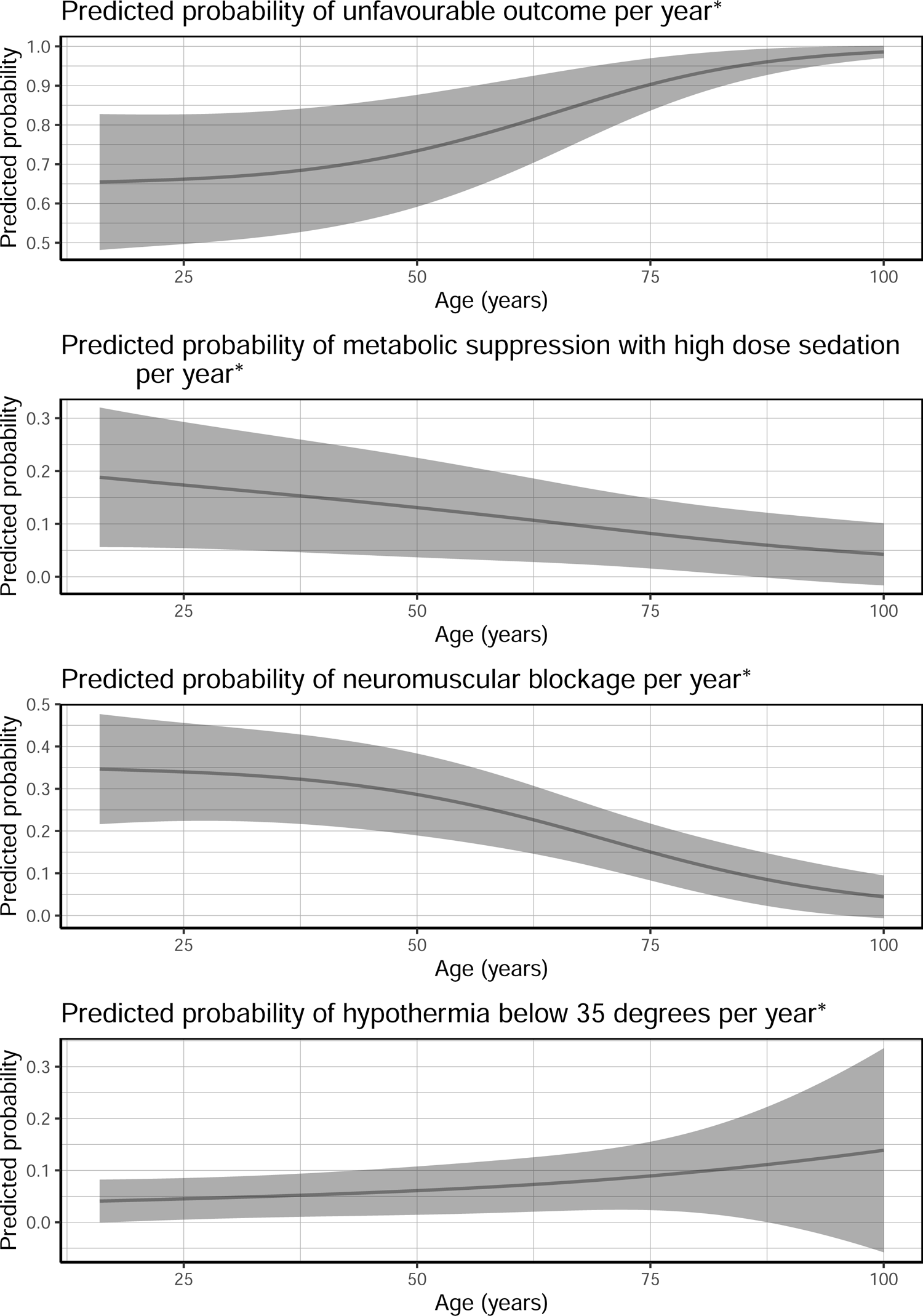
Predicted probabilities of unfavorable outcome, metabolic suppression, neuromuscular blockage and hypothermia

**Figure 3.**
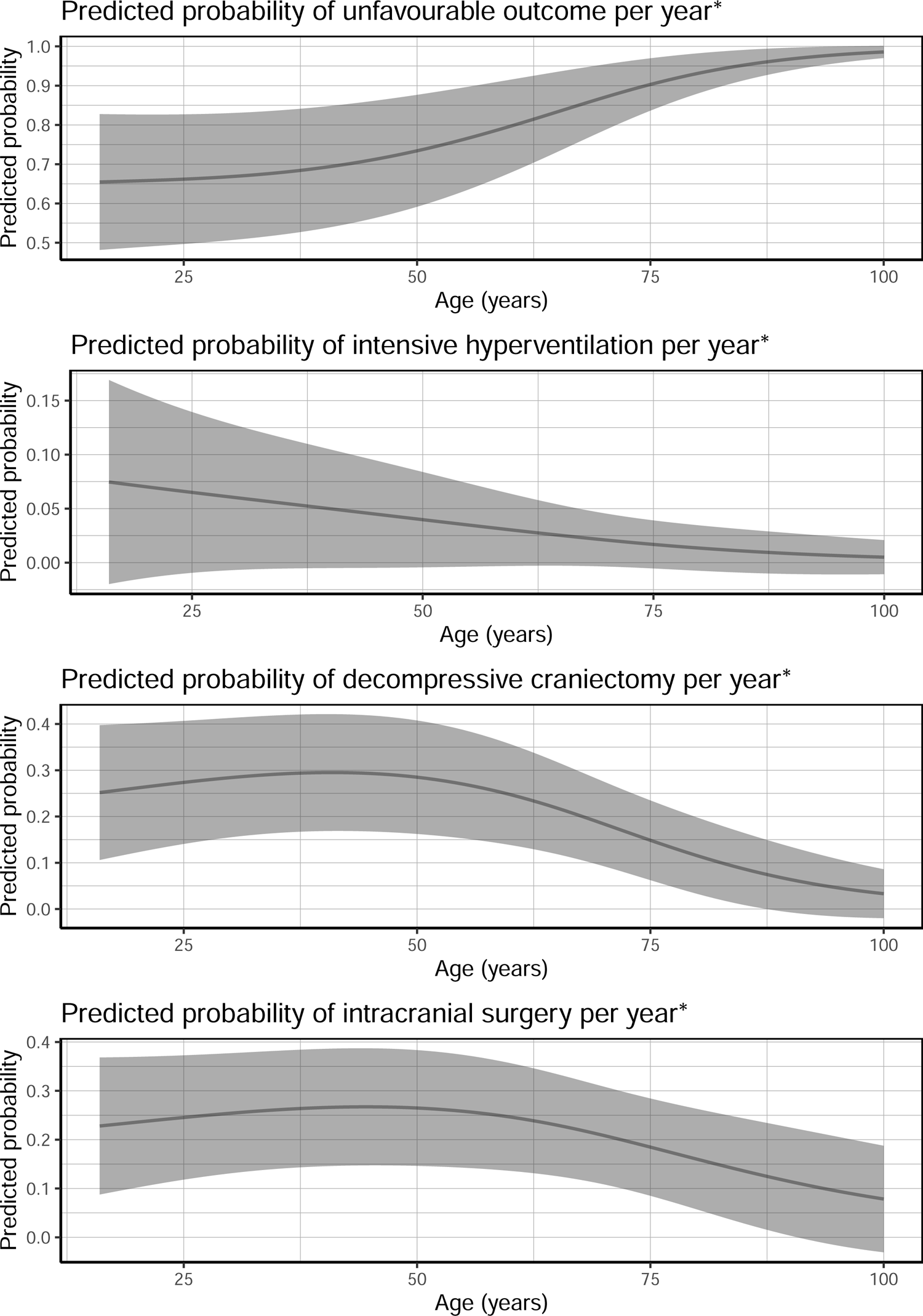
Predicted probabilities of unfavorable outcome, intensive hyperventilation, decompressive craniectomy and intracranial surgery

**Table 3.** Effect of age on outcome and on the probability of receiving one or more high intensity treatments.

|  |  |  |
| --- | --- | --- |
|  | OR* per 10 years [CI] for patients ≤65 | OR* per 10 years [CI] for patients ≥65 |
| **Entire cohort** |  |  |
| GOSE | 0.90 [0.84 - 0.91] | 0.62 [0.54 - 0.70] |
| Emergency intracranial surgery | 0.98 [0.89 - 1.07] | 0.43 [0.32 - 0.56] |
| ICU admission | 0.94 [0.87 - 1.01] | 0.63 [0.52 - 0.78] |
| **ICU cohort** |  |  |
| GOSE | 0.84 [0.80 - 0.91] | 0.48 [0.33 - 0.52] |
| Metabolic suppression | 0.85 [0.76 - 0.93] | 0.90 [0.63 - 1.38] |
| Neuromuscular blockade | 0.88 [0.80 - 0.96] | 0.60 [0.41 - 0.90] |
| Intensive hypoxia | 0.88 [0.66 - 1.08] | 0.22 [0.06 - 0.86] |
| Hypothermia below 35°C | 1.13 [0.95 - 1.34] | 1.27 [0.74 - 2.05] |
| Decompressive craniectomy | 0.99 [0.86 - 1.12] | 0.43 [0.24 - 0.75] |
| Intracranial surgery | 1.05 [0.90 - 1.19] | 0.51 [0.30 - 0.84] |

Abbreviations: CI = confidence interval; GOSE = Glasgow Outcome Scale Extended; OR = Odds Ratio; ICU = intensive care unit
\*An OR <1 indicates a lower probability of having a good outcome or of receiving the particular high intensity treatment per 10 years. For example, the OR of being admitted to the ICU is 0.63 for a 75 year old compared to a 65 year old. The OR per year can be calculated by taking the 10<sup>th</sup> root (10<sup>1/x</sup>) of the displayed OR.
\*\*All models of the entire cohort are adjusted for age; gender; cause of injury (road traffic accident, fall, or other); American Society of Anesthesiologists Physical Status (ASAPS) classification; pre-injury use of anticoagulants or platelet inhibitors; pre-injury use of beta blockers; GCS motor score at baseline; pupillary reactivity at baseline; major extracranial injury (MEI); Marshall CT classification.
\*\*\* All models of the entire cohort are adjusted for age; gender; cause of injury (road traffic accident, fall, or other); American Society of Anesthesiologists Physical Status (ASAPS) classification; pre-injury use of anticoagulants or platelet inhibitors; pre-injury use of beta blockers; GCS motor score at baseline; pupillary reactivity at baseline; major extracranial injury (MEI); Marshall CT classification; median ICP before receiving the first HIT on the ICU.

#### Entire cohort (Table 3-5, Figure 1)

After the age of 65, increasing age was independently associated with a lower probability of receiving emergency intracranial surgery (OR per 10 years = 0.43, CI = 0.32 – 0.56) (Table 3, Figure 1). Further, increasing age was associated with a lower probability of being admitted to the ICU (OR per 10 years if over 65 = 0.63, CI = 0.52 – 0.78) (Table 3, Figure 1). Similar results, were found for patients under the age of 65, for emergency intracranial surgery (OR per 10 years = 0.98, CI = 0.89 – 1.07), and ICU admission (OR per 10 years = 0.94, CI = 0.87 – 1.01) (Table 3). In the sensitivity analyses, comparable results were found (Table 4-5).

**Table 4.** Sensitivity analyses - Effect of age on outcome and on the probability of receiving one or more high intensity treatments.

|  |  |  |
| --- | --- | --- |
|  | OR* per 10 years [CI] for patients ≤50 | OR* per 10 years [CI] for patients ≥50 |
| **Entire cohort\*\*** |  |  |
| GOSE | 0.88 [0.81 - 0.92] | 0.81 [0.73 - 0.84] |
| Emergency intracranial surgery | 1.11 [0.98 - 1.28] | 0.63 [0.53 - 0.71] |
| ICU admission | 0.99 [0.89 - 1.09] | 0.75 [0.67 - 0.86] |
| **ICU cohort\*\*\*** |  |  |
| GOSE | 0.85 [0.80 - 0.95] | 0.67 [0.58 - 0.73] |
| Metabolic suppression | 0.90 [0.77 - 1.01] | 0.79 [0.66 - 0.97] |
| Neuromuscular blockade | 0.95 [0.83 - 1.07] | 0.69 [0.57 - 0.83] |
| Intensive hypoxia | 0.76 [0.51 - 1.02] | 0.80 [0.50 - 1.32] |
| Hypothermia below 35°C | 1.10 [0.85 - 1.41] | 1.22 [0.92 - 1.61] |
| Decompressive craniectomy | 1.07 [0.86 - 1.28] | 0.70 [0.55 - 0.92] |
| Intracranial surgery | 1.04 [0.83 - 1.25] | 0.85 [0.68 - 1.09] |

Abbreviations: CI = confidence interval; GOSE = Glasgow Outcome Scale Extended; OR = Odds Ratio; ICU = intensive care unit
\*An OR <1 indicates a lower probability of having a good outcome or of receiving the particular high intensity treatment per 10 years. For example, the OR of being admitted to the ICU in the entire cohort is 0.75 for a 60 year old compared to a 50 year old. The OR per year can be calculated by taking the 10<sup>th</sup> root (10<sup>1/x</sup>) of the displayed OR.
\*\*All models of the entire cohort are adjusted for age; gender; cause of injury (road traffic accident, fall, or other); American Society of Anesthesiologists Physical Status (ASAPS) classification; pre-injury use of anticoagulants or platelet inhibitors; pre-injury use of beta blockers; GCS motor score at baseline; pupillary reactivity at baseline; major extracranial injury (MEI); Marshall CT classification.
\*\*\* All models of the entire cohort are adjusted for age; gender; cause of injury (road traffic accident, fall, or other); American Society of Anesthesiologists Physical Status (ASAPS) classification; pre-injury use of anticoagulants or platelet inhibitors; pre-injury use of beta blockers; GCS motor score at baseline; pupillary reactivity at baseline; major extracranial injury (MEI); Marshall CT classification; median ICP before receiving the first HIT on the ICU.

**Table 5.** Sensitivity analyses - Effect of age on outcome and on the probability of receiving one or more high intensity treatments.

|  |  |  |
| --- | --- | --- |
|  | OR* per 10 years [CI] for patients ≤80 | OR* per 10 years [CI] for patients ≥80 |
| **Entire cohort\*\*** |  |  |
| GOSE | 0.87 [0.81 - 0.88] | 0.22 [0.18 - 0.43] |
| Emergency intracranial surgery | 0.88 [0.81 - 0.94] | 0.21 [0.06 - 0.58] |
| ICU admission | 0.91 [0.85 - 0.97] | 0.17 [0.08 - 0.37] |
| **ICU cohort\*\*\*** |  |  |
| GOSE | 0.79 [0.75 - 0.84] | 0.11 [0.08 - 0.15] |
| Metabolic suppression | 0.86 [0.79 - 0.93] | 0.44 [0.02 - 2.82] |
| Neuromuscular blockade | 0.84 [0.78 - 0.92] | 0.37 [0.03 - 1.93] |
| Intensive hypoxia | 0.78 [0.63 - 0.97] | 0.00 [0.00 - Inf] |
| Hypothermia below 35°C | 1.17 [1.00 - 1.34] | 0.67 [0.03 - 4.01] |
| Decompressive craniectomy | 0.89 [0.79 - 0.998] | 0.85 [0.06 - 5.95] |
| Intracranial surgery | 0.98 [0.86 - 1.09] | 0.00 [0.00 - 0.27] |

Abbreviations: CI = confidence interval; GOSE = Glasgow Outcome Scale Extended; OR = Odds Ratio; ICU = intensive care unit
\*An OR <1 indicates a lower probability of having a good outcome or of receiving the particular high intensity treatment per 10 years. For example, the OR of being admitted to the ICU is 0.17 for a 90 year old compared to a 80 year old. The OR per year can be calculated by taking the 10<sup>th</sup> root ( $10^{\sqrt{x}}$ ) of the displayed OR.
\*\*All models of the entire cohort are adjusted for age; gender; cause of injury (road traffic accident, fall, or other); American Society of Anesthesiologists Physical Status (ASAPS) classification; pre-injury use of anticoagulants or platelet inhibitors; pre-injury use of beta blockers; GCS motor score at baseline; pupillary reactivity at baseline; major extracranial injury (MEI); Marshall CT classification.
\*\*\* All models of the entire cohort are adjusted for age; gender; cause of injury (road traffic accident, fall, or other); American Society of Anesthesiologists Physical Status (ASAPS) classification; pre-injury use of anticoagulants or platelet inhibitors; pre-injury use of beta blockers; GCS motor score at baseline; pupillary reactivity at baseline; major extracranial injury (MEI); Marshall CT classification; median ICP before receiving the first HIT on the ICU.

#### ICU cohort (Table 3-5, Figure 2-3)

For patients over 65, an increase in age was independently associated with a lower probability of receiving neuromuscular blockade (OR per 10 years = 0.60, CI = 0.41 – 0.90), a lower likelihood of receiving intensive hypocapnia (OR per 10 years if over 65 = 0.22, CI = 0.06 – 0.86), a lower likelihood of decompressive craniectomy (OR per 10 years = 0.43, CI = 0.24 – 0.75), and a lower probability for intracranial surgery (OR per 10 years = 0.51, CI = 0.30 – 0.84). When older than 65, increasing age was not associated with a lower probability of receiving metabolic suppression with high dose barbiturates or propofol (OR per 10 years = 0.90, CI = 0.63 – 1.38), and the likelihood of receiving hypothermic therapy below 35°C (OR per 10 years = 1.27, CI = 0.63 – 1.38) (Table 3, and Figure 1-3). Similar results were found in the sensitivity analyses (Table 4-5).

The graphs show the probability of receiving a certain therapy with increasing age in a patient who did not use anticoagulants or platelet inhibitors, or betablockers, who had a mild systemic disease, whose cause of injury was a fall, after which the GCS motor score was 2, the pupils were both reactive, there were no MEI, and a Marshall CT score of II.

## DISCUSSION

We aimed to confirm the association between age and worse outcome, and to assess the association between age and received high-intensity treatment (HIT). Increasing age is independently associated with a worse outcome, and a lower probability to receive HIT, such as emergency intracranial surgery, ICU admission, and ICP lowering therapies (e.g. intensive hypocapnia, and decompressive craniectomy).

With this new body of evidence, it is important to elucidate that invalid pessimistic assumptions are not the only possible explanation of worse outcome in older patients with TBI. Important factors that may prevent high-intensity treatment for older adults with TBI, include the (previous) wish to withhold treatment of the patient or surrogate, and a higher expected probability of a bad outcome in the opinion of the clinician, patient, or surrogate, based on clinical experience. Furthermore, even though more intensive treatment has been shown to be effective in some older patients with TBI (24–26), pre-existent comorbidities and age are still major drivers of outcome. Focusing on frailty instead of on biological age, may therefore be more relevant. A newly developed, and validated frailty index for patients with TBI, is the CENTER-TBI frailty index score (27). A higher CENTER-TBI frailty index score significantly increases the risk of unfavorable outcome, regardless of age. Clinicians should therefore be reluctant to let age alone drive the decision to apply less intensive treatment, indicating ageism. Ageism can be defined as age-related discrimination (28), and plays a role in all layers of (Western) society (29). The influence of ageism in the hospital setting became more apparent during the covid-19 pandemic (30). Ageism negatively impacts the probability of a positive outcome for older patients (31), potentially fueling a self-fulfilling prophecy.

Multiple previous studies indicate the possible existence of a self-fulfilling prophecy. Skaansar et al. (15) recently warned for the possibility of a self-fulfilling prophecy after finding that treatment intensity decreased with advanced age, and that a decrease in treatment intensity was associated with an increased risk of mortality, concluding that mortality may be lower if HIT would be given to older patients (15). Kirkman et al. (32) found that it took longer for older patients to obtain an initial head CT, that the chances of being transferred to a neurotrauma center were lower, and that the CT was less likely to be reviewed by a senior physician (32). Some centers have suggested age-dependent limitations for more intensive treatments (33–35). Other centers tend to be more liberal and are inclined to admit all older adults after TBI to the ICU for neurological assessments and head CTs (36). Moreover, in the majority of centers in Europe, age played a role in the decision to withdraw LSM (37), and the decision to perform neurosurgical procedures (14). A recent study, however, showed that age was not associated with faster withdrawal of LSM (38). Future research should elucidate if a self-fulfilling prophecy, induced by ageism may be a relevant cause of higher mortality in older patients with TBI. A randomized controlled trial on this topic may be ethically challenging. Therefore, another option is comparative effectiveness research between hospitals, or countries with different treatment regimes. One such option may be a comparison between Chinese centers and European centers, since a provider profiling between different neurotrauma centers suggested that China less often applies restrictions in therapies based on age (39).

We found differences between baseline characteristics in patients over 65 and younger patients. In line with previous literature (40), the GCS motor score on admission generally seemed to be more favorable in patients over 65. Two important explanations may play a role here. One explanation may be the cause of injury. As seen in our study, and also in the study of Lecky et al. (41) on the CENTER-TBI registry, elderly patients more often have a low-energy fall compared with younger patients. Another explanation pertains to atrophy of the brain associated with ageing. Due to this natural process, older patients may have higher intracranial compliance in case of minor or moderate space occupying traumatic lesions, and, consequently, less often need high-intensity treatments to manage ICP. However, given the statistical adjustment for pupillary reactivity and Marshall CT score, we think this possible confounding factor of brain atrophy with higher age should not be relevant for our analyses.

Furthermore, we found that patients older than 65 years more often used beta-blockers, anticoagulants, or platelet inhibitors. Beta-blockers may improve outcomes after TBI in adults (42). The use of anticoagulants is associated with worse initial TBI severity (43–46) and some studies have reported worse outcomes and higher need for neurosurgical intervention (43, 44, 47–49). Moreover, we found more CT abnormalities for older patients, in line with previous literature (50, 51). The higher prevalence in older patients may result from various factors, ranging from biological factors such as vessels being more vulnerable to rupture, more atrophy (leaving more space for blood within the skull), and the higher use of anticoagulants (52–54).

The CENTER-TBI study is unique for its extensive data collection in multiple centers, enrolling TBI patients with varying injury severity across a wide range of European centers. Furthermore, the observational design of the CENTER-TBI study, ensures larger generalizability of the results compared to a clinical trial with strict enrollment criteria (55). There are also limitations that should be considered when interpreting the results. First, all centers in the CENTER-TBI study are committed to TBI research. This might limit generalizability since it is only a sample of the neuro-trauma centers in Europe. Second, HIT was based on clinical considerations that could have differed in a different setting (other country, other clinicians). We included a variety of high-intensity treatments, but cannot guarantee that we included all relevant treatments. However, given the consistency of our results across all therapies in different cohorts, including sensitivity analyses, we expect results to be similar for other treatments. Third, we may not have controlled for all relevant factors that could have influenced the outcome or the treatment decision. Fourth, for statistical reasons, it would have been preferable to have included more patients, especially older patients. Last, there was some missing data. However, to lower the impact of missing data we multiply imputed missing data of variables that were included in our models.

## CONCLUSION

Older patients have poorer outcome, and were less likely to receive high-intensity treatments, independent of patient and injury characteristics. Clinicians should not withhold high-intensity treatments solely based on older age. Educating clinicians about this delicate topic, and performing further comparative effectiveness research focusing on older patients may improve diagnosis, treatments, and understanding of TBI outcomes in this group.

## DECLARATIONS

### Ethics approval and consent to participate

The Medical Ethics Committees of all participating centers approved the CENTER-TBI study, and informed consent was obtained according to local regulations

### Availability of data and materials

The data supporting the findings in the study are available upon reasonable request from the corresponding Author (EvV) and are stored at https://center-tbi.incf.org/

## Authors’ contributions

EvV analyzed the data and drafted the manuscript, and the supplementary tables and figures. All coauthors gave feedback on the manuscript. HL supervised the project. All coauthors gave feedback on (and approved) the final version of the manuscript.

## CENTER-TBI INVESTIGATORS AND PARTICIPANTS

Cecilia Åkerlund1, Krisztina Amrein2, Nada Andelic3, Lasse Andreassen4, Audny Anke5, Anna Antoni6, Gérard Audibert7, Philippe Azouvi8, Maria Luisa Azzolini9, Ronald Bartels10, Pál Barzó11, Romuald Beauvais12, Ronny Beer13, Bo-Michael Bellander14, Antonio Belli15, Habib Benali16, Maurizio Berardino17, Luigi Beretta9, Morten Blaabjerg18, Peter Bragge19, Alexandra Brazinova20, Vibeke Brinck21, Joanne Brooker22, Camilla Brorsson23, Andras Buki24, Monika Bullinger25, Manuel Cabeleira26, Alessio Caccioppola27, Emiliana Calappi 27, Maria Rosa Calvi9, Peter Cameron28, Guillermo Carbayo Lozano29, Marco Carbonara27, Simona Cavallo17, Giorgio Chevallard30, Arturo Chieregato30, Giuseppe Citerio31, 32, Hans Clusmann33, Mark Coburn34, Jonathan Coles35, Jamie D. Cooper36, Marta Correia37, Amra Čović 38, Nicola Curry39, Endre Czeiter24, Marek Czosnyka26, Claire Dahyot Fizelier40, Paul Dark41, Helen Dawes42, Véronique De Keyser43, Vincent Degos16, Francesco Della Corte44, Hugo den Boogert10, Bart Depreitere45, Đula Đilvesi 46, Abhishek Dixit47, Emma Donoghue22, Jens Dreier48, Guy Loup Dulière49, Ari Ercole47, Patrick Esser42, Erzsébet Ezer50, Martin Fabricius51, Valery L. Feigin52, Kelly Foks53, Shirin Frisvold54, Alex Furmanov55, Pablo Gagliardo56, Damien Galanaud16, Dashiell Gantner28, Guoyi Gao57, Pradeep George58, Alexandre Ghuysen59, Lelde Giga60, Ben Glocker61, Jagoš Golubovic46, Pedro A. Gomez 62, Johannes Gratz63, Benjamin Gravesteijn64, Francesca Grossi44, Russell L. Gruen65, Deepak Gupta66, Juanita A. Haagsma64, Iain Haitsma67, Raimund Helbok13, Eirik Helseth68, Lindsay Horton 69, Jilske Huijben64, Peter J. Hutchinson70, Bram Jacobs71, Stefan Jankowski72, Mike Jarrett21, Ji yao Jiang58, Faye Johnson73, Kelly Jones52, Mladen Karan46, Angelos G. Kolias70, Erwin Kompanje74, Daniel Kondziella51, Evgenios Kornaropoulos47, Lars Owe Koskinen75, Noémi Kovács76, Ana Kowark77, Alfonso Lagares62, Linda Lanyon58, Steven Laureys78, Fiona Lecky79, 80, Didier Ledoux78, Rolf Lefering81, Valerie Legrand82, Aurelie Lejeune83, Leon Levi84, Roger Lightfoot85, Hester Lingsma64, Andrew I.R. Maas43, Ana M. Castaño León62, Marc Maegele86, Marek Majdan20, Alex Manara87, Geoffrey Manley88, Costanza Martino89, Hugues Maréchal49, Julia Mattern90, Catherine McMahon91, Béla Melegh92, David Menon47, Tomas Menovsky43, Ana Mikolic64, Benoit Misset78, Visakh Muraleedharan58, Lynnette Murray28, Ancuta Negru93, David Nelson1, Virginia Newcombe47, Daan Nieboer64, József Nyirádi2, Otesile Olubukola79, Matej Oresic94, Fabrizio Ortolano27, Aarno Palotie95, 96, 97, Paul M. Parizel98, Jean François Payen99, Natascha Perera12, Vincent Perlbarg16, Paolo Persona100, Wilco Peul101, Anna Piippo-Karjalainen102, Matti Pirinen95, Dana Pisica64, Horia Ples93, Suzanne Polinder64, Inigo Pomposo29, Jussi P. Posti 103, Louis Puybasset104, Andreea Radoi 105, Arminas Ragauskas106, Rahul Raj102, Malinka Rambadagalla107, Isabel Retel Helmrich64, Jonathan Rhodes108, Sylvia Richardson109, Sophie Richter47, Samuli Ripatti95, Saulius Rocka106, Cecilie Roe110, Olav Roise111,112, Jonathan Rosand113, Jeffrey V. Rosenfeld114, Christina Rosenlund115, Guy Rosenthal55, Rolf Rossaint77, Sandra Rossi100, Daniel Rueckert61 Martin Rusnák116, Juan Sahuquillo105, Oliver Sakowitz90, 117, Renan Sanchez Porras117, Janos Sandor118, Nadine Schäfer81, Silke Schmidt119, Herbert Schoechl120, Guus Schoonman121, Rico Frederik Schou122, Elisabeth Schwendenwein6, Charlie Sewalt64, Toril Skandsen123, 124, Peter Smielewski26, Abayomi Sorinola125, Emmanuel Stamatakis47, Simon Stanworth39, Robert Stevens126, William Stewart127, Ewout W. Steyerberg64, 128, Nino Stocchetti129, Nina Sundström130, Riikka Takala131, Viktória Tamás125, Tomas Tamosuitis132, Mark Steven Taylor20, Braden Te Ao52, Olli Tenovuo103, Alice Theadom52, Matt Thomas87, Dick Tibboel133, Marjolein Timmers74, Christos Tolias134, Tony Trapani28, Cristina Maria Tudora93, Andreas Unterberg90, Peter Vajkoczy 135, Shirley Vallance28, Egils Valeinis60, Zoltán Vámos50, Mathieu van der Jagt136, Gregory Van der Steen43, Joukje van der Naalt71, Jeroen T.J.M. van Dijck 101, Thomas A. van Essen101, Wim Van Hecke137, Caroline van Heugten138, Dominique Van Praag139, Ernest van Veen64, Thijs Vande Vyvere137, Roel P. J. van Wijk101, Alessia Vargiolu32, Emmanuel Vega83, Kimberley Velt64, Jan Verheyden137, Paul M. Vespa140, Anne Vik123, 141, Rimantas Vilcinis132, Victor Volovici67, Nicole von Steinbüchel38, Daphne Voormolen64, Petar Vulekovic46, Kevin K.W. Wang142, Eveline Wiegers64, Guy Williams47, Lindsay Wilson69, Stefan Winzeck47, Stefan Wolf143, Zhihui Yang113, Peter Ylén144, Alexander Younsi90, Frederick A. Zeiler47,145, Veronika Zelinkova20, Agate Ziverte60, Tommaso Zoerle27

1. Department of Physiology and Pharmacology, Section of Perioperative Medicine and Intensive Care, Karolinska Institutet, Stockholm, Sweden
2. János Szentágothai Research Centre, University of Pécs, Pécs, Hungary
3. Division of Surgery and Clinical Neuroscience, Department of Physical Medicine and Rehabilitation, Oslo University Hospital and University of Oslo, Oslo, Norway
4. Department of Neurosurgery, University Hospital Northern Norway, Tromso, Norway
5. Department of Physical Medicine and Rehabilitation, University Hospital Northern Norway, Tromso, Norway
6. Trauma Surgery, Medical University Vienna, Vienna, Austria
7. Department of Anesthesiology & Intensive Care, University Hospital Nancy, Nancy, France
8. Raymond Poincare hospital, Assistance Publique – Hopitaux de Paris, Paris, France
9. Department of Anesthesiology & Intensive Care, S Raffaele University Hospital, Milan, Italy
10. Department of Neurosurgery, Radboud University Medical Center, Nijmegen, The Netherlands
11. Department of Neurosurgery, University of Szeged, Szeged, Hungary
12. International Projects Management, ARTTIC, Munchen, Germany
13. Department of Neurology, Neurological Intensive Care Unit, Medical University of Innsbruck, Innsbruck, Austria
14. Department of Neurosurgery & Anesthesia & intensive care medicine, Karolinska University Hospital, Stockholm, Sweden
15. NIHR Surgical Reconstruction and Microbiology Research Centre, Birmingham, UK
16. Anesthesie-Réanimation, Assistance Publique – Hopitaux de Paris, Paris, France
17. Department of Anesthesia & ICU, AOU Città della Salute e della Scienza di Torino - Orthopedic and Trauma Center, Torino, Italy
18. Department of Neurology, Odense University Hospital, Odense, Denmark
19. BehaviourWorks Australia, Monash Sustainability Institute, Monash University, Victoria, Australia
20. Department of Public Health, Faculty of Health Sciences and Social Work, Trnava University, Trnava, Slovakia
21. Quesgen Systems Inc., Burlingame, California, USA
22. Australian & New Zealand Intensive Care Research Centre, Department of Epidemiology and Preventive Medicine, School of Public Health and Preventive Medicine, Monash University, Melbourne, Australia
23. Department of Surgery and Perioperative Science, Umeå University, Umeå, Sweden
24. Department of Neurosurgery, Medical School, University of Pécs, Hungary and Neurotrauma Research Group, János Szentágothai Research Centre, University of Pécs, Hungary
25. Department of Medical Psychology, Universitätsklinikum Hamburg-Eppendorf, Hamburg, Germany
26. Brain Physics Lab, Division of Neurosurgery, Dept of Clinical Neurosciences, University of Cambridge, Addenbrooke’s Hospital, Cambridge, UK
27. Neuro ICU, Fondazione IRCCS Cà Granda Ospedale Maggiore Policlinico, Milan, Italy
28. ANZIC Research Centre, Monash University, Department of Epidemiology and Preventive Medicine, Melbourne, Victoria, Australia
29. Department of Neurosurgery, Hospital of Cruces, Bilbao, Spain
30. NeuroIntensive Care, Niguarda Hospital, Milan, Italy
31. School of Medicine and Surgery, Università Milano Bicocca, Milano, Italy
32. NeuroIntensive Care, ASST di Monza, Monza, Italy
33. of Neurosurgery, Medical Faculty RWTH Aachen University, Aachen, Germany
34. Department of Anesthesiology and Intensive Care Medicine, University Hospital Bonn, Bonn, Germany
35. Department of Anesthesia & Neurointensive Care, Cambridge University Hospital NHS Foundation Trust, Cambridge, UK
36. School of Public Health & PM, Monash University and The Alfred Hospital, Melbourne, Victoria, Australia
37. Radiology/MRI department, MRC Cognition and Brain Sciences Unit, Cambridge, UK
38. Institute of Medical Psychology and Medical Sociology, Universitätsmedizin Göttingen, Göttingen, Germany
39. Oxford University Hospitals NHS Trust, Oxford, UK
40. Intensive Care Unit, CHU Poitiers, Potiers, France
41. University of Manchester NIHR Biomedical Research Centre, Critical Care Directorate, Salford Royal Hospital NHS Foundation Trust, Salford, UK
42. Movement Science Group, Faculty of Health and Life Sciences, Oxford Brookes University, Oxford, UK
43. Department of Neurosurgery, Antwerp University Hospital and University of Antwerp, Edegem, Belgium
44. Department of Anesthesia & Intensive Care, Maggiore Della Carità Hospital, Novara, Italy
45. Department of Neurosurgery, University Hospitals Leuven, Leuven, Belgium
46. Department of Neurosurgery, Clinical centre of Vojvodina, Faculty of Medicine, University of Novi Sad, Novi Sad, Serbia
47. Division of Anaesthesia, University of Cambridge, Addenbrooke’s Hospital, Cambridge, UK
48. Center for Stroke Research Berlin, Charité – Universitätsmedizin Berlin, corporate member of Freie Universität Berlin, Humboldt-Universität zu Berlin, and Berlin Institute of Health, Berlin, Germany
49. Intensive Care Unit, CHR Citadelle, Liège, Belgium
50. Department of Anaesthesiology and Intensive Therapy, University of Pécs, Pécs, Hungary
51. Departments of Neurology, Clinical Neurophysiology and Neuroanesthesiology, Region Hovedstaden Rigshospitalet, Copenhagen, Denmark
52. National Institute for Stroke and Applied Neurosciences, Faculty of Health and Environmental Studies, Auckland University of Technology, Auckland, New Zealand
53. Department of Neurology, Erasmus MC, Rotterdam, the Netherlands
54. Department of Anesthesiology and Intensive care, University Hospital Northern Norway, Tromso, Norway
55. Department of Neurosurgery, Hadassah-hebrew University Medical center, Jerusalem, Israel
56. Fundación Instituto Valenciano de Neurorrehabilitación (FIVAN), Valencia, Spain
57. Department of Neurosurgery, Shanghai Renji hospital, Shanghai Jiaotong University/school of medicine, Shanghai, China
58. Karolinska Institutet, INCF International Neuroinformatics Coordinating Facility, Stockholm, Sweden
59. Emergency Department, CHU, Liège, Belgium
60. Neurosurgery clinic, Pauls Stradins Clinical University Hospital, Riga, Latvia
61. Department of Computing, Imperial College London, London, UK
62. Department of Neurosurgery, Hospital Universitario 12 de Octubre, Madrid, Spain
63. Department of Anesthesia, Critical Care and Pain Medicine, Medical University of Vienna, Austria
64. Department of Public Health, Erasmus Medical Center-University Medical Center, Rotterdam, The Netherlands
65. College of Health and Medicine, Australian National University, Canberra, Australia
66. Department of Neurosurgery, Neurosciences Centre & JPN Apex trauma centre, All India Institute of Medical Sciences, New Delhi-110029, India
67. Department of Neurosurgery, Erasmus MC, Rotterdam, the Netherlands
68. Department of Neurosurgery, Oslo University Hospital, Oslo, Norway
69. Division of Psychology, University of Stirling, Stirling, UK
70. Division of Neurosurgery, Department of Clinical Neurosciences, Addenbrooke’s Hospital & University of Cambridge, Cambridge, UK
71. Department of Neurology, University of Groningen, University Medical Center Groningen, Groningen, Netherlands
72. Neurointensive Care, Sheffield Teaching Hospitals NHS Foundation Trust, Sheffield, UK
73. Salford Royal Hospital NHS Foundation Trust Acute Research Delivery Team, Salford, UK
74. Department of Intensive Care and Department of Ethics and Philosophy of Medicine, Erasmus Medical Center, Rotterdam, The Netherlands
75. Department of Clinical Neuroscience, Neurosurgery, Umeå University, Umeå, Sweden
76. Hungarian Brain Research Program - Grant No. KTIA_13_NAP-A-II/8, University of Pécs, Pécs, Hungary
77. Department of Anaesthesiology, University Hospital of Aachen, Aachen, Germany
78. Cyclotron Research Center, University of Liège, Liège, Belgium
79. Centre for Urgent and Emergency Care Research (CURE), Health Services Research Section, School of Health and Related Research (ScHARR), University of Sheffield, Sheffield, UK
80. Emergency Department, Salford Royal Hospital, Salford UK
81. Institute of Research in Operative Medicine (IFOM), Witten/Herdecke University, Cologne, Germany
82. VP Global Project Management CNS, ICON, Paris, France
83. Department of Anesthesiology-Intensive Care, Lille University Hospital, Lille, France
84. Department of Neurosurgery, Rambam Medical Center, Haifa, Israel
85. Department of Anesthesiology & Intensive Care, University Hospitals Southhampton NHS Trust, Southhampton, UK
86. Cologne-Merheim Medical Center (CMMC), Department of Traumatology, Orthopedic Surgery and Sportmedicine, Witten/Herdecke University, Cologne, Germany
87. Intensive Care Unit, Southmead Hospital, Bristol, Bristol, UK
88. Department of Neurological Surgery, University of California, San Francisco, California, USA
89. Department of Anesthesia & Intensive Care,M. Bufalini Hospital, Cesena, Italy
90. Department of Neurosurgery, University Hospital Heidelberg, Heidelberg, Germany
91. Department of Neurosurgery, The Walton centre NHS Foundation Trust, Liverpool, UK
92. Department of Medical Genetics, University of Pécs, Pécs, Hungary
93. Department of Neurosurgery, Emergency County Hospital Timisoara, Timisoara, Romania
94. School of Medical Sciences, Örebro University, Örebro, Sweden
95. Institute for Molecular Medicine Finland, University of Helsinki, Helsinki, Finland
96. Analytic and Translational Genetics Unit, Department of Medicine; Psychiatric & Neurodevelopmental Genetics Unit, Department of Psychiatry; Department of Neurology, Massachusetts General Hospital, Boston, MA, USA
97. Program in Medical and Population Genetics; The Stanley Center for Psychiatric Research, The Broad Institute of MIT and Harvard, Cambridge, MA, USA
98. Department of Radiology, University of Antwerp, Edegem, Belgium
99. Department of Anesthesiology & Intensive Care, University Hospital of Grenoble, Grenoble, France
100. Department of Anesthesia & Intensive Care, Azienda Ospedaliera Università di Padova, Padova, Italy
101. Dept. of Neurosurgery, Leiden University Medical Center, Leiden, The Netherlands and Dept. of Neurosurgery, Medical Center Haaglanden, The Hague, The Netherlands
102. Department of Neurosurgery, Helsinki University Central Hospital
103. Division of Clinical Neurosciences, Department of Neurosurgery and Turku Brain Injury Centre, Turku University Hospital and University of Turku, Turku, Finland
104. Department of Anesthesiology and Critical Care, Pitié -Salpêtrière Teaching Hospital, Assistance Publique, Hôpitaux de Paris and University Pierre et Marie Curie, Paris, France
105. Neurotraumatology and Neurosurgery Research Unit (UNINN), Vall d’Hebron Research Institute, Barcelona, Spain
106. Department of Neurosurgery, Kaunas University of technology and Vilnius University, Vilnius, Lithuania
107. Department of Neurosurgery, Rezekne Hospital, Latvia
108. Department of Anaesthesia, Critical Care & Pain Medicine NHS Lothian & University of Edinburg, Edinburgh, UK
109. Director, MRC Biostatistics Unit, Cambridge Institute of Public Health, Cambridge, UK
110. Department of Physical Medicine and Rehabilitation, Oslo University Hospital/University of Oslo, Oslo, Norway
111. Division of Orthopedics, Oslo University Hospital, Oslo, Norway
112. Institue of Clinical Medicine, Faculty of Medicine, University of Oslo, Oslo, Norway
113. Broad Institute, Cambridge MA Harvard Medical School, Boston MA, Massachusetts General Hospital, Boston MA, USA
114. National Trauma Research Institute, The Alfred Hospital, Monash University, Melbourne, Victoria, Australia
115. Department of Neurosurgery, Odense University Hospital, Odense, Denmark
116. International Neurotrauma Research Organisation, Vienna, Austria
117. Klinik für Neurochirurgie, Klinikum Ludwigsburg, Ludwigsburg, Germany
118. Division of Biostatistics and Epidemiology, Department of Preventive Medicine, University of Debrecen, Debrecen, Hungary
119. Department Health and Prevention, University Greifswald, Greifswald, Germany
120. Department of Anaesthesiology and Intensive Care, AUVA Trauma Hospital, Salzburg, Austria
121. Department of Neurology, Elisabeth-TweeSteden Ziekenhuis, Tilburg, the Netherlands
122. Department of Neuroanesthesia and Neurointensive Care, Odense University Hospital, Odense, Denmark
123. Department of Neuromedicine and Movement Science, Norwegian University of Science and Technology, NTNU, Trondheim, Norway
124. Department of Physical Medicine and Rehabilitation, St.Olavs Hospital, Trondheim University Hospital, Trondheim, Norway
125. Department of Neurosurgery, University of Pécs, Pécs, Hungary
126. Division of Neuroscience Critical Care, John Hopkins University School of Medicine, Baltimore, USA
127. Department of Neuropathology, Queen Elizabeth University Hospital and University of Glasgow, Glasgow, UK
128. Dept. of Department of Biomedical Data Sciences, Leiden University Medical Center, Leiden, The Netherlands
129. Department of Pathophysiology and Transplantation, Milan University, and Neuroscience ICU, Fondazione IRCCS Cà Granda Ospedale Maggiore Policlinico, Milano, Italy
130. Department of Radiation Sciences, Biomedical Engineering, Umeå University, Umeå, Sweden
131. Perioperative Services, Intensive Care Medicine and Pain Management, Turku University Hospital and University of Turku, Turku, Finland
132. Department of Neurosurgery, Kaunas University of Health Sciences, Kaunas, Lithuania
133. Intensive Care and Department of Pediatric Surgery, Erasmus Medical Center, Sophia Children’s Hospital, Rotterdam, The Netherlands
134. Department of Neurosurgery, Kings college London, London, UK
135. Neurologie, Neurochirurgie und Psychiatrie, Charité – Universitätsmedizin Berlin, Berlin, Germany
136. Department of Intensive Care Adults, Erasmus MC– University Medical Center Rotterdam, Rotterdam, the Netherlands
137. icoMetrix NV, Leuven, Belgium
138. Movement Science Group, Faculty of Health and Life Sciences, Oxford Brookes University, Oxford, UK
139. Psychology Department, Antwerp University Hospital, Edegem, Belgium
140. Director of Neurocritical Care, University of California, Los Angeles, USA
141. Department of Neurosurgery, St.Olavs Hospital, Trondheim University Hospital, Trondheim, Norway
142. Department of Emergency Medicine, University of Florida, Gainesville, Florida, USA
143. Department of Neurosurgery, Charité – Universitätsmedizin Berlin, corporate member of Freie Universität Berlin, Humboldt-Universität zu Berlin, and Berlin Institute of Health, Berlin, Germany
144. VTT Technical Research Centre, Tampere, Finland
145. Section of Neurosurgery, Department of Surgery, Rady Faculty of Health Sciences, University of Manitoba, Winnipeg, MB, Canada

